# Predicting the potential impact of scaling up four pneumonia interventions on under-five pneumonia mortality: A prospective LiST analysis for Bangladesh, Chad, and Ethiopia

**DOI:** 10.1101/2023.04.19.23288780

**Authors:** Theresa Pfurtscheller, Felix Lam, Rasheduzzaman Shah, Rana Shohel, Maria Suau Sans, Narcisse Tounaikok, Abas Hassen, Alemayehu Berhanu, Dinkineh Bikila, Elizabeth Berryman, Tedila Habte, Leith Greenslade, Rebecca Nantanda, Kevin Baker

## Abstract

Pneumonia remains the leading cause of mortality in children under five outside the neonatal period. Progress has slowed down in the last decade and increased efforts to scale-up effective pneumonia interventions are needed.

We used the Lives Saved Tool (LiST), a modelling software for child mortality in low- and middle-income settings, to prospectively analyse the potential impact of upscaling pneumonia interventions in Bangladesh, Chad, and Ethiopia from 2023-2030. Haemophilus influenzae type B (Hib) vaccination, pneumococcal conjugate vaccine (PCV), oral antibiotics, pulse oximetry and oxygen were included as pneumonia interventions in our analysis. Outcomes were number of pneumonia deaths averted, proportion of deaths averted by intervention, and changes in the under-five mortality rate.

Our results show that 19,775 lives of children under five could be saved in Bangladesh, 76,470 in Chad, and 97,343 in Ethiopia by scaling intervention coverages to ≥90% by 2030. Our estimated reductions in pneumonia deaths among children under five range from 44.61% to 57.91% in the respective countries. Increased coverage of oral antibiotics, pulse oximetry, and oxygen show similar effects in all three countries, averting between 18.80% and 23.65% of expected pneumonia deaths. Scaling-up PCV has a prominent effect, especially in Chad where it could avert 14.04% of expected pneumonia deaths. Under-five mortality could be reduced by 1.42/1000, live births in Bangladesh, 22.52/1000, in Chad, and 5.48/1000, in Ethiopia.

This analysis shows the high impact of upscaling pneumonia interventions. The lack of data regarding coverage indicators is a barrier for further research, as well as policy and implementation, all requiring increased attention.

**Lay summary:** Pneumonia remains the leading cause of death in children under five after the first 28 days of live. However, progress in decreasing pneumonia deaths has stagnated in the worst-affected regions.

This study used a modelling software called the Lives Saved Tool (LiST) to project pneumonia deaths and the number of lives saved in children under five in Bangladesh, Chad, and Ethiopia if four key pneumonia interventions (vaccinations, oral antibiotic treatment, pulse oximetry, and oxygen treatment) were scaled up to a coverage ≥ 90% by 2030.

Our results show that from 2023 to 2030 19,775 lives of children under five with pneumonia could be saved in Bangladesh, 76,470 in Chad, and 97,343 in Ethiopia. Increasing oral antibiotics, pulse oximetry, and oxygen coverages proved highly valuable for reducing pneumonia deaths in all three countries. Pneumococcal vaccination had an especially prominent effect in Chad.

Our analysis shows the potential of the four interventions for improving child health in high burden pneumonia countries and highlights the importance of increased funding to reduce childhood pneumonia. The lack of up-to-date accurate data, especially for pulse oximetry and oxygen coverage indicators, is a barrier not only for research but also for evidence-based policy-making that needs to be addressed.

## Background

Pneumonia remains the leading cause of morbidity and mortality in children under five outside of the neonatal period in low- and middle-income countries (LMICs) (1). Globally, approximately 700,000 children die of pneumonia each year before reaching the age of five, with South-Asia and western and central Africa disproportionately affected (2). While we have seen dramatic reductions in the prevalence of acute respiratory infections (ARI) from 1990-2010, progress has significantly slowed down in the last decade, especially in western and central Africa, where pneumonia prevalence rates have stagnated (3). To reach the Sustainable Development Goals (SDGs) globally, interventions that are known to be effective need to be scaled up and innovations to further reduce the pneumonia burden are needed in the most affected regions (4, 5).

Preventive interventions such as vaccines and risk factor reduction played a key role in pneumonia control in the past (4). The haemophilus influenza B (Hib) vaccine and pneumococcal conjugate vaccines (PCV) have reduced pneumonia deaths and cases, thus contributing to increased survival for children in low-income environments (6-10). Evidence suggests that PCV has a greater impact on health outcomes in low-income environments compared to middle- and high-income environments (11). However, PCV uptake is still low in many high-burden pneumonia countries, and some countries, such as Chad, have yet to introduce PCV nationally (12, 13). Besides increased vaccine coverage, new diagnostic support is needed to further reduce pneumonia mortality in children (1, 14). Hypoxaemia, defined as blood oxygen saturation (SpO2) of <90%, is highly prevalent in children with respiratory symptoms and is associated with higher mortality (15, 16). Pulse oximeters can identify those children at higher odds of death by measuring SpO2 (15) thus facilitating a purposeful and targeted use of oxygen treatment (1). The combined use of pulse oximetry and oxygen has been shown to effectively decrease pneumonia mortality in low resource settings, when used by frontline health workers (17-20). Supportive care such as oxygen has become increasingly important as major causes of pneumonia shift from bacterial to viral pathogens in LMICs and PCV and Hib vaccine coverages increase (21). However, both oxygen and pulse oximetry are not commonly available in these settings (22-24). Oral and parenteral antibiotics remain key components of pneumonia treatment in children (1). While in many high- and middle-income settings antimicrobial resistance and the over and inappropriate use of antibiotics are problematic, low-income settings such as Bangladesh, Chad, and Ethiopia have concerningly low coverages of oral antibiotics for childhood pneumonia (25-27).

There are implementation and policy gaps in effective preventive and curative pneumonia interventions (28). Increased efforts to effectively use known and new interventions are much needed, especially in high burden countries.

Prospective estimations of the effects scaled intervention coverages can have in specific settings can guide policy makers in their decision making and prioritization of certain interventions in the face of resource constraints. The Lives Saved Tool (LiST), initially developed at Johns Hopkins Bloomberg School of Public Health, has frequently been used to inform child and maternal health policy decisions by modelling effects of potential future scenarios or through evaluating the impact of implemented interventions (29-33). A coverage indicator of pulse oximetry and oxygen has recently been added into the LiST model based on new effectiveness estimates (19). Following this addition, our study aims to provide impact estimates of scaling-up four pneumonia interventions on national pneumonia mortality reductions and the number of children’s lives saved during the SDG era in three high burden pneumonia countries Bangladesh, Chad, and Ethiopia.

## Methodology

### Study design

This study used LiST to perform a prospective analysis of the potential impact of upscaling four pneumonia interventions in Bangladesh, Chad, and Ethiopia on pneumonia-specific mortality and under five mortality rates (U5MR) from 2023 to 2030. The interventions included in our analysis were: Hib vaccine, PCV, oral antibiotics, pulse oximetry, and oxygen. These interventions were selected as they are the primary interventions in LiST that are directly linked to pneumonia morbidity and mortality, rather than affecting outcomes through changing risk factors (e.g. reducing stunting).

### Study setting

Bangladesh, Chad, and Ethiopia have been selected as case studies for this study due to their high burden of childhood pneumonia and their different demographic, economic, and social profiles. While Bangladesh and Ethiopia are highly populous countries, Chad’s population is considerably smaller. Further, both Ethiopia’s and Chad’s populations are more dispersed than Bangladesh’s dense population (34, 35). The Sub-Saharan settings of Chad and Ethiopia are classified as low-income (36). Bangladesh, situated in South Asia is a lower-middle income setting (36).

Table 1 summarizes baseline health indicators and coverage rates of key pneumonia interventions in these three countries. Bangladesh’s U5MR at 29.11 per 1000 live births is fairly close to the SDG goal of 25/1000, live births, whereas Chad’s mortality rate in children under five remains high at 110/1000, live births despite the progress made in the past decades (37). Ethiopia had been one of the few countries to achieve Millennium Development Goal 4 in 2015 (38). However, in order to reach the SDG goal 3.2, the country needs to decrease its current U5MR of 48.71 by almost 50% (37).

**Table 1.** Baseline Health Indicators and Pneumonia intervention coverage rates Bangladesh, Chad, Ethiopia

|  | Bangladesh | Chad | Ethiopia |
| --- | --- | --- | --- |
| U5MR (37) | 29.1 | 110.05 | 48.71 |
| Number of deaths from pneumonia in children under five (60) | 7,031.24 | 14,709.98 | 18,558.56 |
| % of post-neonatal deaths in children under five by pneumonia (60, 61) | 16.66% | 28.05% | 23.63% |
| HiB vaccine coverage (13) | 98% | 58% | 65% |
| PCV coverage (13) | 99% | 0% | 61% |
| Oral antibiotics for pneumonia coverage□ | 34% (25) | 17.6% (26) | 32% (27) |
| Pulse oximetry (Pox) and oxygen for pneumonia coverage | 3.17 % (62) | 0.69 % □ (48) | 11.25% □ |

□ approximated by number of children with ARI symptoms taken to a health care provider
□ approximated through pulse oximeter/oxygen availability and care seeking behaviour for ARI

**Table 2.**
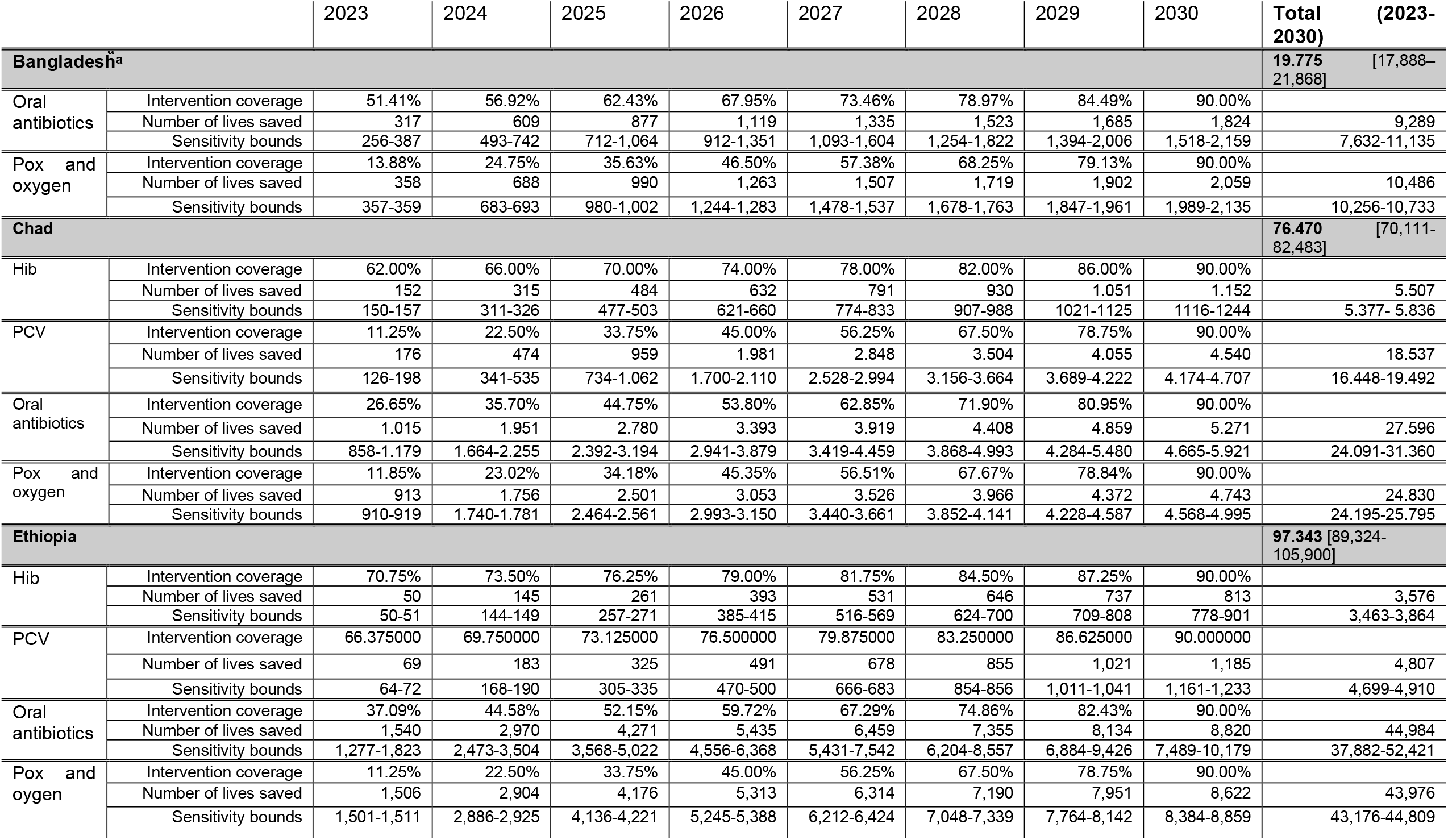
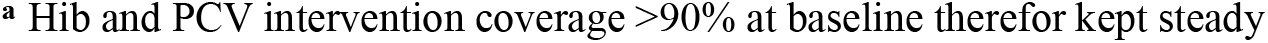
Projected coverage estimates and number of lives saved by intervention, country, and year

|  |  | 2023 | 2024 | 2025 | 2026 | 2027 | 2028 | 2029 | 2030 | Total (2023-2030) |
| --- | --- | --- | --- | --- | --- | --- | --- | --- | --- | --- |
| **Bangladesh** □ □ □ □ □ □ □ □ □ □ |  |  |  |  |  |  |  |  |  | **19.775** [17,888–21,868] |
| Oral antibiotics | Intervention coverage | 51.41% | 56.92% | 62.43% | 67.95% | 73.46% | 78.97% | 84.49% | 90.00% |  |
|  | Number of lives saved | 317 | 609 | 877 | 1,119 | 1,335 | 1,523 | 1,685 | 1,824 | 9,289 |
|  | Sensitivity bounds | 256-387 | 493-742 | 712-1,064 | 912-1,351 | 1,093-1,604 | 1,254-1,822 | 1,394-2,006 | 1,518-2,159 | 7,632-11,135 |
| Pox and oxygen | Intervention coverage | 13.88% | 24.75% | 35.63% | 46.50% | 57.38% | 68.25% | 79.13% | 90.00% |  |
|  | Number of lives saved | 358 | 688 | 990 | 1,263 | 1,507 | 1,719 | 1,902 | 2,059 | 10,486 |
|  | Sensitivity bounds | 357-359 | 683-693 | 980-1,002 | 1,244-1,283 | 1,478-1,537 | 1,678-1,763 | 1,847-1,961 | 1,989-2,135 | 10,256-10,733 |
| **Chad** |  |  |  |  |  |  |  |  |  | **76.470** [70,111-82,483] |
| Hib | Intervention coverage | 62.00% | 66.00% | 70.00% | 74.00% | 78.00% | 82.00% | 86.00% | 90.00% |  |
|  | Number of lives saved | 152 | 315 | 484 | 632 | 791 | 930 | 1,051 | 1,152 | 5,507 |
|  | Sensitivity bounds | 150-157 | 311-326 | 477-503 | 621-660 | 774-833 | 907-988 | 1021-1125 | 1116-1244 | 5,377- 5,836 |
| PCV | Intervention coverage | 11.25% | 22.50% | 33.75% | 45.00% | 56.25% | 67.50% | 78.75% | 90.00% |  |
|  | Number of lives saved | 176 | 474 | 959 | 1,981 | 2,848 | 3,504 | 4,055 | 4,540 | 18,537 |
|  | Sensitivity bounds | 126-198 | 341-535 | 734-1,062 | 1,700-2,110 | 2,528-2,994 | 3,156-3,664 | 3,689-4,222 | 4,174-4,707 | 16,448-19,492 |
| Oral antibiotics | Intervention coverage | 26.65% | 35.70% | 44.75% | 53.80% | 62.85% | 71.90% | 80.95% | 90.00% |  |
|  | Number of lives saved | 1,015 | 1,951 | 2,780 | 3,393 | 3,919 | 4,408 | 4,859 | 5,271 | 27,596 |
|  | Sensitivity bounds | 858-1,179 | 1,664-2,255 | 2,392-3,194 | 2,941-3,879 | 3,419-4,459 | 3,868-4,993 | 4,284-5,480 | 4,665-5,921 | 24,091-31,360 |
| Pox and oxygen | Intervention coverage | 11.85% | 23.02% | 34.18% | 45.35% | 56.51% | 67.67% | 78.84% | 90.00% |  |
|  | Number of lives saved | 913 | 1,756 | 2,501 | 3,053 | 3,526 | 3,966 | 4,372 | 4,743 | 24,830 |
|  | Sensitivity bounds | 910-919 | 1,740-1,781 | 2,464-2,561 | 2,993-3,150 | 3,440-3,661 | 3,852-4,141 | 4,228-4,587 | 4,568-4,995 | 24,195-25,795 |
| **Ethiopia** |  |  |  |  |  |  |  |  |  | **97.343** [89,324-105,900] |
| Hib | Intervention coverage | 70.75% | 73.50% | 76.25% | 79.00% | 81.75% | 84.50% | 87.25% | 90.00% |  |
|  | Number of lives saved | 50 | 145 | 261 | 393 | 531 | 646 | 737 | 813 | 3,576 |
|  | Sensitivity bounds | 50-51 | 144-149 | 257-271 | 385-415 | 516-569 | 624-700 | 709-808 | 778-901 | 3,463-3,864 |
| PCV | Intervention coverage | 66.375000 | 69.750000 | 73.125000 | 76.500000 | 79.875000 | 83.250000 | 86.625000 | 90.000000 |  |
|  | Number of lives saved | 69 | 183 | 325 | 491 | 678 | 855 | 1,021 | 1,185 | 4,807 |
|  | Sensitivity bounds | 64-72 | 168-190 | 305-335 | 470-500 | 666-683 | 854-856 | 1,011-1,041 | 1,161-1,233 | 4,699-4,910 |
| Oral antibiotics | Intervention coverage | 37.09% | 44.58% | 52.15% | 59.72% | 67.29% | 74.86% | 82.43% | 90.00% |  |
|  | Number of lives saved | 1,540 | 2,970 | 4,271 | 5,435 | 6,459 | 7,355 | 8,134 | 8,820 | 44,984 |
|  | Sensitivity bounds | 1,277-1,823 | 2,473-3,504 | 3,568-5,022 | 4,556-6,368 | 5,431-7,542 | 6,204-8,557 | 6,884-9,426 | 7,489-10,179 | 37,882-52,421 |
| Pox and oxygen | Intervention coverage | 11.25% | 22.50% | 33.75% | 45.00% | 56.25% | 67.50% | 78.75% | 90.00% |  |
|  | Number of lives saved | 1,506 | 2,904 | 4,176 | 5,313 | 6,314 | 7,190 | 7,951 | 8,622 | 43,976 |
|  | Sensitivity bounds | 1,501-1,511 | 2,886-2,925 | 4,136-4,221 | 5,245-5,388 | 6,212-6,424 | 7,048-7,339 | 7,764-8,142 | 8,384-8,859 | 43,176-44,809 |

□ Hib and PCV intervention coverage >90% at baseline therefore kept steady

Coverage of key pneumonia interventions varies between the settings. Bangladesh reports full coverage (defined as >90%) of Hib vaccine and PCV whereas estimates for Ethiopia show room for improvement and Chad has not yet introduced PCV into its national vaccine program (13). The coverage of oral antibiotics as a key intervention for childhood pneumonia can be considered low (<35%) in all three countries (25-27). Ethiopia has a national oxygen and pulse oximetry roadmap which has helped to provide oxygen in all hospitals and 26% of health centres (39). In Bangladesh pulse oximetry was only available in 6% of public health facilities in 2018 (40). Oxygen was more widely available with 27% of district hospitals reporting an oxygen source. However, at lower levels of the health system oxygen availability was 18% or less (40). Specific estimates for pulse oximetry and oxygen availability in Chad were not available, several national documents however, indicated highly fragmented availability of oxygen sources in questionable maintenance (41-43).

### Analysis

LiST is a modelling software that can estimate the impact of upscaled intervention coverages on child and maternal mortality (44). The deterministic, linear, mathematical modelling tool describes fixed relations between inputs and outputs (44). The methodology of LiST has been described in more detail elsewhere (44). We included four interventions in our analysis based on their link to pneumonia mortality within the LiST software (45). We chose not to include preventive zinc supplementation as an intervention in our analysis, even though this intervention is linked to pneumonia mortality within LiST. This was based on the low quality of evidence and lack of statistical significance of the reductions in pneumonia-specific mortality through preventive zinc supplementation in the literature (46). We also did not include interventions indirectly linked to pneumonia through risk factors, such as wasting, stunting, or indoor air pollution. Target levels for 2030 were set to an aspirational full coverage level, defined as ≥90%, for all countries and annual values were linearly interpolated from estimated baseline values in 2022 to 2030. Interventions that showed coverages of or above 90% at baseline were kept steady. All estimations are presented with their sensitivity bounds, which are obtained by LiST through multiplication of lower and upper bound intervention effectiveness estimates with the number of potentially averted deaths (47).

### Data Sources

Besides defined target coverage levels, LiST requires baseline intervention coverages and health status measures, as well as estimates of intervention effectiveness (44). The software uses automatic data inputs from various data sources for these indicators and provides the user with a list of these. We manually reviewed all data inputs on baseline health status and intervention coverages and replaced them with more recent data where available.

For this process we used the latest Demographic Health Surveys (DHS), Multiple Indicator Cluster Surveys (MICS), WHO and UNICEF vaccination coverage estimates (WUENIC), the WHO mortality data base, the Global Burden of Disease Study 2019, peer reviewed publications, and national data retrieved from the district health information system 2 (DHIS2) to establish an up-to-date, credible baseline data set. We used U5MR estimates from the United Nations Interagency Group for Child Mortality for 2020 to estimate baseline mortality.

Coverage data for pulse oximetry and oxygen for childhood pneumonia was largely lacking. While reports about the availability of pulse oximeters, or an oxygen source are more easily available they do not provide an accurate estimate of the coverage of pulse oximetry and oxygen for pneumonia. It needs to be considered that appropriate care at a health facility is often not sought for children with pneumonia symptoms. In Bangladesh we therefore used the proportion of children assessed in public health facilities that had an oxygen saturation measured and recorded in DHIS2 to approximate coverage of pulse oximetry and oxygen for children with pneumonia. For Chad, while national reports from the Ministry of Health and the WHO Chad indicated very fragmented coverage (42, 43), no national estimate could be found. To approximate coverage we have therefore used a regional estimate of oxygen availability for Sub-Saharan Africa derived from multiple Service Provision Assessment surveys (48), multiplied with a national estimate of the proportion of children with ARI for whom care at a health facility was sought (26). For Ethiopia, we used a similar process and multiplied the proportion of children with ARI for whom care at a health facility was sought with the proportion of health facilities with pulse oximeters and oxygen available. As composite indicators for both oxygen and pulse oximetry availability were not accessible, we had to use a pulse oximetry specific indicator for Bangladesh, and an oxygen specific indicator for Chad to approximate the coverage of both interventions together. A summary of all indicators and respective data sources used in our analysis can be found in the supplementary material. Table 1 shows the baseline coverage data in 2022 for the four interventions included in our prospective analysis. We used LiST’s default intervention effectiveness measures (8, 19, 49, 50).

### Outcomes

The primary outcomes of this study were reductions in pneumonia deaths per country, year, and number of lives saved per country, year, and intervention. The reduction in U5MR per country was a secondary outcome.

## Results

### Number of pneumonia deaths and estimated lives saved

Figure 1.a-1c show the expected number of pneumonia deaths between 2023 and 2030 if all interventions were linearly scaled up to 90% versus no scale-up (all intervention coverages remain at baseline levels). In Bangladesh, under-five pneumonia deaths decline from 5,784 in 2022 to 1,401 (sensitivity bounds: 1,137–1,632) by 2030 due to increased intervention coverage. In the counterfactual, with intervention coverages remaining flat at 2022 levels, under-5 pneumonia deaths also decline, though at a slower rate from 5,784 deaths in 2022 to 5,284 by 2030. As a result, we estimate that 19,775 (sensitivity bounds: 17,888–21,868) lives could be saved between 2023-2030 due to intervention coverage scale-up. This represents 44.61% of pneumonia deaths that would be expected between 2023-2030 without increased coverage.

**Figure 1.**
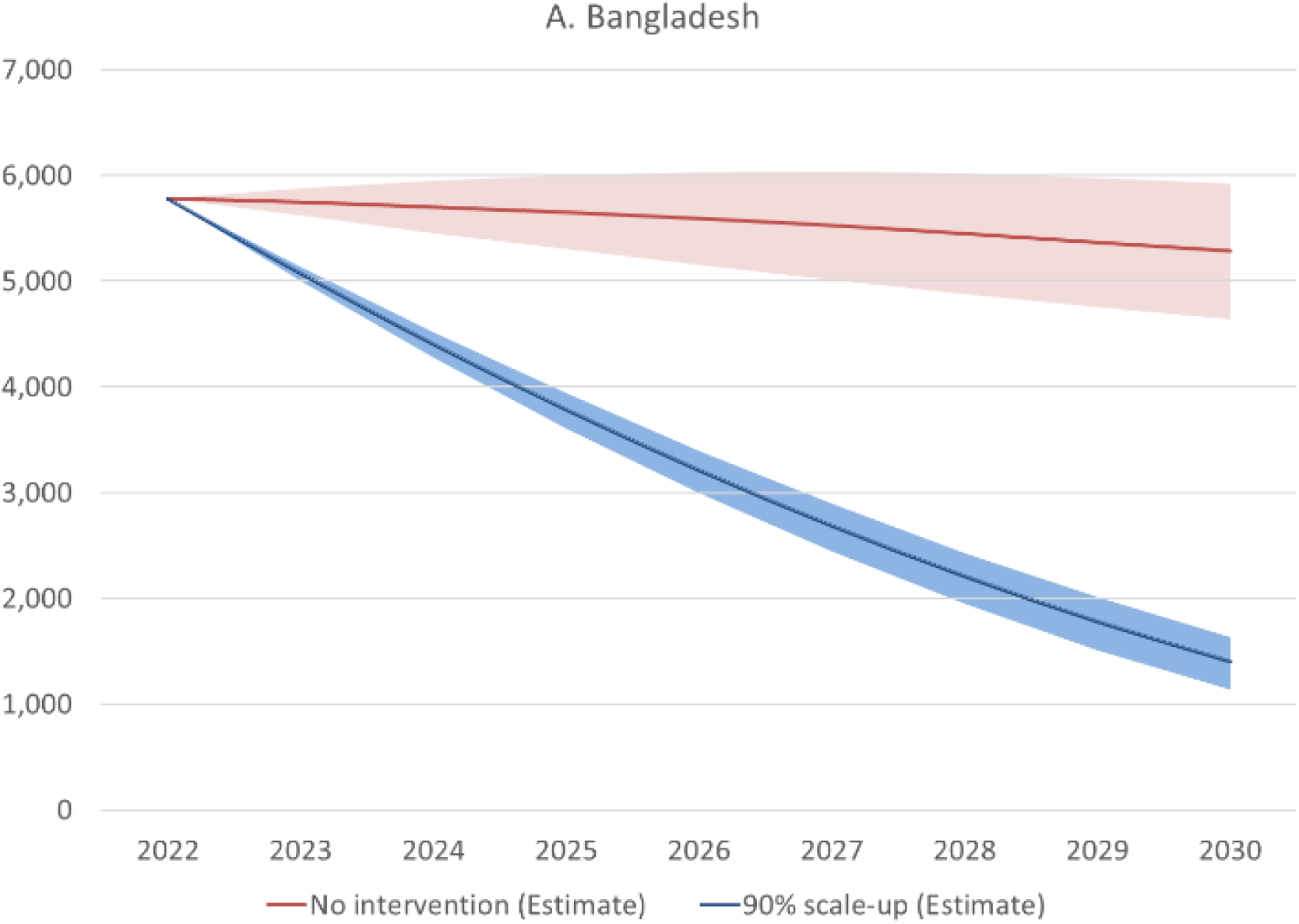
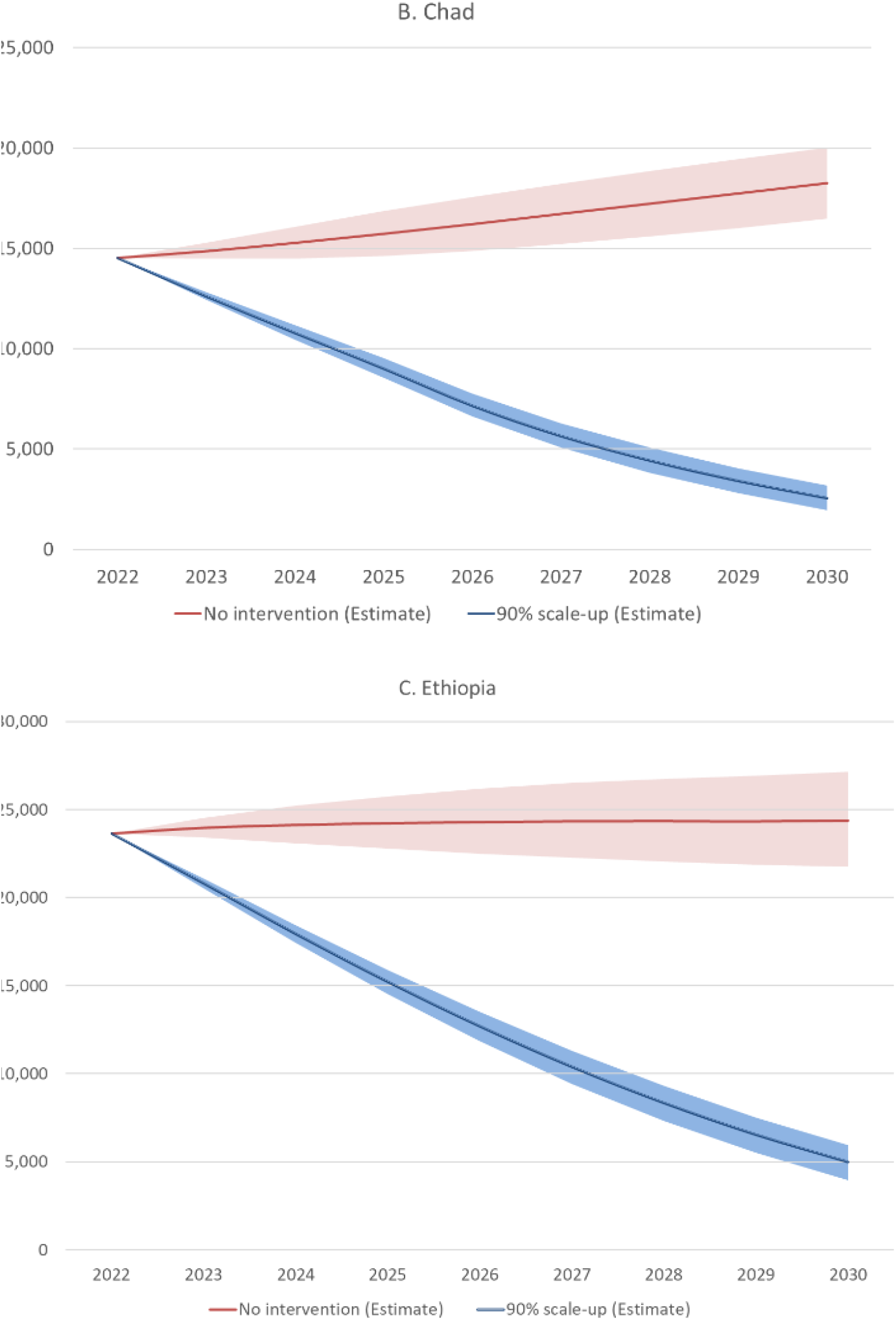
Estimated pneumonia deaths with 90% scale-up of inteventions versus no scale-up, 2022-2030

In Chad, under-5 pneumonia deaths are projected to decline from 14,537 to 2,543 (sensitivity bounds: 1,945–3,166) with scale-up of interventions as compared to the counterfactual where under-5 pneumonia deaths are projected to increase from 14,537 to 18,249 with no change in intervention coverages. We estimate that 76,470 (sensitivity bounds: 70,111-82,483) lives would be saved between 2023-2030, thus averting 57.91% of expected pneumonia deaths.

In Ethiopia, pneumonia deaths decline from 23,647 in 2022 to 4,999 by 2030 (sensitivity bounds: 3,940-5,956) with intervention scale-up. Without any increases in intervention coverage, pneumonia deaths are expected to increase from 23,647 to 24,393. In Ethiopia scaled coverage of four interventions has the potential to save 97,343 lives (sensitivity bounds: 89,324-105,900), thus averting 50.15% of expected pneumonia deaths. including annual projected coverage levels. 5,507 saved lives, 7.20% of all potential lives saved from 2023-2030 in Chad are attributable to scaled Hib vaccination coverage. In Ethiopia scaled Hib vaccination coverage contributes to 3,576, 3.67% of all potential lives saved. PCV introduction and scale-up contributes to 5,507 lives saved (24.24% of total) in Chad and 3,576 (4.94% of total) in Ethiopia. Increasing the coverage of oral antibiotics from 2023-2030 could save 9,289 (46.97% of total) lives in Bangladesh, 27,596 (36.09% of total) in Chad, and 44,984 (46.21% of total) in Ethiopia. Increasing coverage of pulse oximetry and oxygen for childhood pneumonia could save 10,486 (53,03% of total) lives in Bangladesh, 24,830 (32.47% of total) in Chad, and 43,976 (45.18% of total) in Ethiopia. Figure 2 provides a graphical representation of the proportional shares of lives saved by each intervention for the 2023-2030 period for each country.

**Figure 2.**
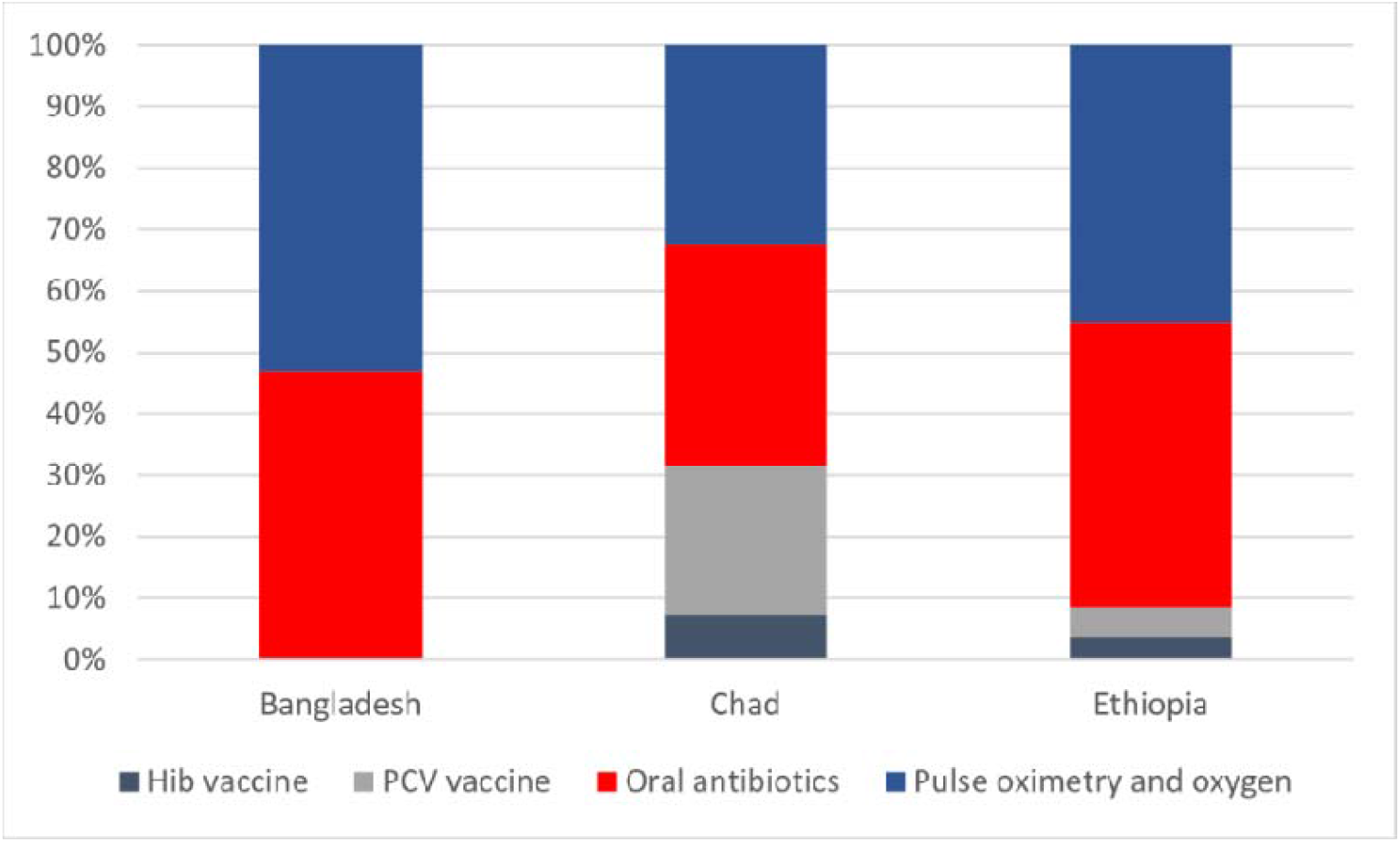
Proportion of total lives saved between 2022-2030 by intervention

### Reductions in under-five mortality rates

In Bangladesh our results indicate a reduction in U5MR attributable to the four investigated pneumonia interventions of 1.42 per 1000 live births by 2030. In Chad our results show a decline in U5MR by 22.52 deaths per 1000, live births. For Ethiopia U5MR could decrease by 5.48 deaths per 1000, live births in 2030 if the four included pneumonia interventions would be scaled up.

## Discussion

In all three countries, around half of the expected pneumonia deaths could be averted if the key pneumonia interventions included in our analysis were scaled up to ≥90% coverage between 2023 and 2030. The continuity of these results indicates the value of the four pneumonia interventions included in our analysis across settings and different baseline health conditions of a population.

Our results show that increased coverage of oral antibiotics for pneumonia could play a major role in reducing childhood pneumonia mortality. This trend is in line with past reports of antibiotic treatment as a key strategy to reduce pneumonia mortality in children (51). The result also specifically highlights that this intervention has not yet been optimised. The potential benefit of available antibiotic treatment for childhood pneumonia however can only be realized if care seeking at health facilities is simultaneously strengthened. Healthcare-seeking has been described as especially low for respiratory infections in children and if care is sought, informal providers are often the first contact point for caregivers which can lead to an ineffective and inappropriate use of antibiotics (52, 53).

Our results show a similar potential for pulse oximetry and oxygen in reducing pneumonia mortality. Our search for data points of this variable confirms the low availability of this intervention in low-income settings in Sub-Saharan Africa and South Asia (22-24), despite pulse oximeters being considered easy to use by frontline health workers (54). Consequently, the high potential of both pulse oximetry and oxygen is virtually unused in high burden contexts that require accelerated progress to reach SDG 3.2. The same considerations regarding care seeking at health facilities mentioned above apply to pulse oximetry and oxygen as an intervention to reduce pneumonia mortality. Additionally, referral mechanisms and referral adherence are highly important to reach the full potential of pulse oximetry and oxygen as this intervention aims to filter out severely ill children at lower health care levels and treat them accordingly in higher level referral hospitals (55, 56). Following the experiences of the COVID-19 pandemic increasing oxygen availability in settings where currently oxygen treatment is rarely used should be linked to capacity building (57) and the lessons learnt during the pandemic could be transferred to severe childhood pneumonia. Additional funding is needed in low- and middle-income settings to realize the potential this intervention could have for children’s lives, but also to cover the rising treatment costs resulting from increased care seeking behaviour and improved diagnosis of severe illness necessitating escalated treatment (58). Alongside increased funding, further research is required, both at intervention and health system level, to understand barriers to optimising these interventions at scale, as highlighted in recent pneumonia research priorities (28).

The results on potential lives saved through increased coverage of two vaccine interventions affecting pneumonia mortality, Hib vaccination and PCV, are more diverse within the three investigated settings and their effect should be seen in context with their baseline coverage in the respective countries. PCV uptake and scale-up in Chad could avert a considerable proportion of expected deaths. This result highlights the importance for the country to start implementing PCV within its national vaccine schedule. The effects of Hib and PCV vaccine interventions in Ethiopia is not as prominent as that of PCV in Chad; this should however be seen in light of our outcome definition as additional lives saved. Thus, the continued contribution that an already high vaccine coverage has on pneumonia mortality is not depicted in our results. Nevertheless, high or full vaccine coverages should be maintained as they are contributing substantially to decreasing pneumonia mortality, thus providing a more favourable baseline for other interventions.

Looking at the effect the combined scale-up of four pneumonia interventions has on U5MR in the three countries we can see a meaningful impact reducing child mortality in all countries, thus bringing them closer to the SDG 3.2 target of 25 deaths per 1000, live births. In Chad this potential decrease is hugely important for a country that was estimated to have the third highest U5MR worldwide in 2020 (37). In Ethiopia which, which is currently projected to achieve SDG 3.2 targets by 2033 (37), an additional decrease in U5MR of 5.48 deaths per 1000, live births within the next seven years could help the country continue its success from the Millennium Development Goal era and achieve SDG 3.2 before the deadline. In Bangladesh increasing only those four interventions could account for more than a third of the reduction the country needs to achieve SDG 3.2.

More accurate estimates of intervention coverages are greatly needed. Although estimates of national vaccine coverage data are routinely published jointly by the WHO and UNICEF, there is a severe lack of coverage data for antibiotics, pulse oximetry, and oxygen. Due to methodological challenges in gathering antibiotic coverage for pneumonia using household surveys, we settled on using care seeking for symptoms of pneumonia as a proxy. Robust and comparable data on pulse oximetry and oxygen were even more challenging to find across these three countries. Pulse oximetry use for screening children with pneumonia for hypoxemia is reported nationally in Bangladesh’s health management information system. However, data on the prevalence of hypoxemia amongst those screened and coverage of oxygen treatment for those identified with hypoxemia are not. In Ethiopia, estimates of availability of pulse oximeters and oxygen at health facilities are available. The availability of these services at facilities does not however reflect the coverage of these interventions for children with pneumonia. Therefore, we applied a discount factor using the proportion of pneumonia cases seeking care at facilities. In Chad, we were unable to find any data related to pulse oximetery and oxygen, and therefore had to rely on regional estimates from secondary analyses of health facility data to estimate coverage. Lack of data on intervention coverage (particularly antibiotics, pulse oximetry, and oxygen) limits the accuracy of our analysis, but most importantly, it hinders efforts to improve access to these interventions by hiding the severity of the issue. Strengthening national data systems to collect, analyse, and report on intervention coverage and other measures of intervention access is greatly needed.

In our analysis, we forecast coverage of pneumonia interventions to demonstrate their potential impact on reducing mortality and hold constant other interventions. However, it is likely that coverage of other interventions will change over the period as well. There are ongoing efforts to improve neonatal survival, nutrition outcomes, access to family planning, and so forth. Changes in coverage of these interventions may affect our estimates by changing the population of children under five susceptible to pneumonia in our model, adjusting the overall mortality rate or proportion of mortality attributable to pneumonia, and modify risk factors related to pneumonia and child mortality. We examined options to forecast other intervention coverage levels as well. The primary aim of this study though was to better understand the relative impact of specifically upscaling different pneumonia interventions under varying contexts for the purpose of priority setting. For this we compromised on the objective of accurately forecasting to reductions in child deaths related to advances in other child health interventions.

## Conclusion

Pneumonia, the major cause of death in children 1-59 months old in low- and middle income settings (1), does not receive the corresponding global attention in terms of funding or policy engagement (59). Through the example of three high burden countries our analysis shows the effect increased investments and efforts in pneumonia interventions could have in a variety of settings. The lack of data regarding coverage indicators of pneumonia interventions, as well as baseline health indicators related to childhood mortality is a barrier for research, as well as national and international programs that needs to be urgently addressed in order to enhance the validity of future research and enable informed policy making based on strong evidence.

## Data Availability

All data produced in the present study are available upon reasonable request to the authors.

